# Approbation of the Keen Eye Computer-Based Method for Diagnosing Visual Neglect

**DOI:** 10.1101/2025.04.23.25326114

**Authors:** Elizaveta Vasyura, Maria Kovyazina, Georgiy Stepanov, Olga Russkikh, Daria Terentiy, Victoria Propustina, Anatoliy Skvortsov, Nataliya Varako, Svetlana Vasilyeva, Vadim Daminov, Yuri Zinchenko

**Affiliations:** Faculty of Psychology, Lomonosov Moscow State University, Moscow, Russia; Federal Scientific Center of Psychological and Multidisciplinary Research, Moscow, Russia; Research Center of Neurology, Moscow, Russia; State Autonomous Healthcare Institution of the Perm Region “City Clinical Hospital № 4”, Perm, Russia; Academician Ye. A. Vagner Perm State Medical University of the Ministry of Healthcare of the Russian Federation, Perm, Russia; Medical Rehabilitation Clinic, Pirogov National Medical and Surgical Center, Moscow, Russia

**Author notes:** Corresponding author (EV).

## Abstract

This study focuses on the approbation of the Keen Eye method for diagnosing visual neglect. A total of 102 patients with right hemisphere damage participated in the study, 38 of whom were diagnosed with neglect. The primary goal was to assess the effectiveness of Keen Eye in comparison with traditional paper-based tests. Keen Eye is based on a dual-task paradigm, integrating target identification and visual stimulus localization tasks, with stimuli presented in different areas of the screen. The results demonstrated the high diagnostic accuracy of the method, enabling the detection of covert forms of neglect. The study showed that the method allows for the assessment of both horizontal and vertical neglect severity and the identification of patients with neglect combined with non-lateralized attention deficits. The method exhibited high criterion and construct convergent validity and holds significant potential for clinical application.

## Introduction

### Neglect Syndrome

Unilateral spatial neglect is a heterogeneous and multi-component syndrome [1] characterized by the lack of conscious perception of stimuli in the space contralateral to the brain lesion, as well as by non-lateralized attention deficits [2]. Neglect syndrome is a common consequence of right hemisphere damage, with prevalence rates ranging from 43% to 82% among patients in the acute phase of stroke [3–6]. Neither gender nor handedness significantly affects the likelihood of developing neglect following right hemisphere damage [4]. Neglect syndrome manifests across various sensory modalities [1], with visual neglect (VN) being the most functionally limiting for patients. VN significantly impairs daily functioning, affecting essential activities such as eating, personal hygiene, and mobility [7].

Left-sided neglect is a significant negative prognostic factor for post-brain injury functioning. It is reliably associated with poor functional recovery outcomes, and research has shown that the severity of VN serves as an independent predictor of lower recovery outcomes in daily activities after stroke [8, 9]. In everyday life, VN has a substantial negative impact on independence in daily activities—patients tend to ignore the left side while reading, writing, navigating, and working [10].

VN following right hemisphere damage is more common than neglect in other sensory modalities [11]. While neglect can manifest in visual, tactile, and auditory modalities—either in isolated or combined forms—its severity is often greater in the visual domain [11, 12]. This disproportionate severity may be related to the more significant role of automatic attentional processes toward ipsilateral stimuli in the visual modality compared to tactile and auditory modalities.

VN can manifest in both the horizontal and vertical planes, affecting patients’ perception of near and far space [13]. The primary symptoms of VN have been predominantly described in the horizontal plane as an inability to attend to stimuli on the left side of space [14]. When performing visuospatial tests, patients may fail to notice targets on the left side, deviate to the right when bisecting lines, and omit the left side of drawings [15].

Vertical neglect often coexists with horizontal neglect [16, 17]. This condition manifests as inattention to stimuli in the upper or lower parts of the visual field. When combined with horizontal neglect, it frequently results in neglect of the lower left portion of the visual field [16, 18]. Vertical neglect may also be associated with radial neglect, affecting perception in near or far peripersonal space. Studies have shown that patients with VN are more likely to ignore objects in the lower far space [19]. This phenomenon can be explained by differences in spatial reference frames, with errors in radial and vertical bisection likely stemming from impairments in the retinotopic reference system [20]. Vertical neglect can significantly impact daily activities, such as wheelchair navigation and stair climbing, underscoring the importance of its clinical assessment and treatment [21].

In contemporary neglect classification, a subtype of neglect known as vertical neglect is described [22]. Studies have shown that patients with VN tend to ignore the lower portion of the visual field, as evidenced by multiple studies utilizing visual search and line bisection tasks. For example, one study found that 13 patients with VN following a right hemisphere stroke detected fewer targets and made more fixations in the lower-left quadrant [23]. Similar results were obtained from line cancellation tasks in 23 patients with right-sided stroke and experiments using central cues, where more pronounced deficits were observed in the lower part of the visual field [24]. Deviations upwards were often observed in vertical line bisection tests, indicating a lack of attention to lower stimuli [25]. In one case, a 72-year-old patient who had experienced a stroke showed an upward deviation, while in another case, quadrant-specific retinotopic defects persisted irrespective of the direction of gaze [26]. These defects are thought to be linked to disruptions in attention networks, with dysfunction in the dorsal attention network associated with vertical neglect, and damage to the right temporal lobe linked to neglect in the upward direction [25].

### Diagnosis of Visual Neglect

Currently, more than 90 different tests are available for identifying neglect [27]. However, systematic studies indicate that most of these methods lack sufficient validity and reliability [28]. The primary diagnostic approaches include questionnaires, behavioral tests, and traditional paper-and-pencil methods.

Traditional paper-and-pencil tests include cancellation tasks, line bisection tasks, and drawing tests. Among these, the most widely used are the Bell’s Test [29] and letter cancellation tasks, which have demonstrated high accuracy in detecting signs of spatial inattention. However, the line bisection test has limited sensitivity and may fail to identify up to 40% of VN cases [30].

Despite their widespread use, paper-and-pencil methods have low ecological validity, limiting their predictive power for real-world functioning. Additionally, milder forms of VN often go undetected in these tests, and diagnosis may only be possible through the analysis of indirect indicators, such as a right-to-left visual scanning strategy [31].

To address these limitations, more ecologically valid assessment tools have been developed. Among them, the Catherine Bergego Scale (CBS) stands out for its reliability, validity, and sensitivity to changes during rehabilitation [32]. Furthermore, functional tests, computer-based programs, and virtual reality technologies have proven to be more effective in detecting VN compared to paper-and-pencil tests [33, 34].

Given the advantages and limitations of different diagnostic approaches, a combined diagnostic strategy is recommended, incorporating various types of neglect assessments. A comprehensive diagnostic approach integrates traditional paper-and-pencil tests, computer-based tools, and behavioral assessments. This allows for a more precise identification of VN subtypes while ensuring high ecological validity for both clinical practice and research [35, 36]. Thus, modern approaches to VN diagnosis aim for standardization and improved validity, which is particularly important for enhancing diagnostic accuracy and developing effective rehabilitation strategies.

### Dual-Task Paradigm

A modern approach to the diagnosis of VN, particularly its mild forms, involves creating conditions of increased cognitive load to reveal hidden forms of inattention. One such approach is the dual-task paradigm, which encompasses a broad class of methods requiring the simultaneous performance of two tasks—a combination of cognitive/motor, motor/motor, or cognitive/cognitive tasks [37]. The dual-task paradigm is applicable to the study of attention, executive functions [37] working memory [38], and problem-solving [39]. Performing two tasks simultaneously typically leads to interference effects, reflecting the increased demands on the information-processing system [40]. This paradigm has shown promise in detecting mild cognitive impairments and as a rehabilitation tool [37].

Performing two or more tasks simultaneously, especially when each of the parallel tasks is not highly automated, can lead to cognitive overload. A phenomenon known as “attentional blindness” has been described in the context of visual search, occurring when individuals fail to notice unexpected stimuli while engaged in complex tasks [41]. Various neural indicators of different aspects of attentional blindness have been identified: parieto-occipital potentials reflect deficits in target detection, while frontoparietal coherence signals deficits in discrimination [42]. Numerous studies have reported decreased activity in the temporoparietal regions under working memory overload [43–45]. Attentional blindness also modulates activity in the primary visual cortex, leading to reduced responses in visual areas V3 and, possibly, V2 and V1, accompanied by decreased activation in the inferior parietal cortex [46]. Additionally, multitasking affects the awareness of distractors: the higher the load on the selective attention system, the more irrelevant information is ignored [47, 48].

Another related phenomenon is **“**attentional blink”—a temporary reduction in attention following the detection of a visual target [49]. This occurs when processing a large amount of rapidly incoming information. Attentional blink is observed when two stimuli are presented in quick succession, typically within 200–500 ms [50, 51]. Experiments have demonstrated this phenomenon: if stimuli are presented at high speed in the foveal or peripheral visual field, perception of the target stimulus may be impaired if it appears immediately after the preceding one [52, 53]. From a neural perspective, attentional blink involves the right temporoparietal junction (rTPJ) and the left inferior frontal junction (lIFJ) [54].

The right hemisphere plays a particularly significant role in attentional blink. Studies on split-brain patients have shown that when the second target was presented to the right hemisphere, attentional blink lasted significantly longer, indicating the key role of the right hemisphere in processing visual information under limited capacity conditions [55]. In patients with right-hemisphere stroke, the effective field of vision is reduced, and the attentional blink interval is prolonged, particularly for left-sided stimuli [56]. Additionally, transcranial magnetic stimulation (TMS) of the right posterior parietal cortex reduces the magnitude of attentional blink, further confirming the critical role of the right hemisphere in the temporal aspects of visual attention [57].

The right hemisphere is critically important for multitasking in various contexts. Studies have shown that lesions in the right temporoparietal region can lead to chronic multitasking impairments, even after the initial deficits, such as neglect, have been rehabilitated [58]. Neuroimaging research has identified a predominantly right-lateralized frontoparietal network and the cerebellum as key regions activated during dual-task performance [59]. A meta-analysis has revealed a multitasking-related neural network that includes the bilateral intraparietal sulcus, left dorsal premotor cortex, and right anterior insula [60]. However, multitasking and task switching have distinct neural correlates: task switching activates the left premotor and inferior parietal areas, whereas multitasking engages the right prefrontal and inferior parietal cortex [59].

Thus, the limitations of simultaneous and successive cognitive task performance are natural even in a healthy brain, but brain pathology exacerbates the depletion of cognitive resources. It is suggested that cognitive reserve helps protect against the consequences of brain damage. However, the effectiveness of cognitive reserve decreases under multitasking conditions, exposing a “gray zone” between hidden and overt behavioral deficits [62].

The dual-task paradigm is widely used to assess and rehabilitate VN in post-stroke patients. Studies have demonstrated that dual-task conditions, which increase attentional load, can exacerbate VN symptoms and reveal subtle, well-compensated forms of dysfunction [63–65]. Research has also shown that computer-based tasks requiring higher attentional demands are more sensitive than traditional paper-and-pencil tests in detecting VN, as they are less affected by compensatory strategies often employed by patients during recovery [66].

Computerized dual-task assessments lead to more contralesional omissions [66, 67] and slower contralesional reaction times [68] than single-task conditions. Increased cognitive load can reveal left-sided spatial neglect, bilateral extinction, and even vertical extinction in chronic patients [69]. Additionally, large-screen dual-task presentations have been effective in detecting subclinical forms of VN [64]. VN symptoms that remain hidden during single-task conditions may emerge under multitasking demands [62].

Dual-task conditions are not only useful for diagnosing VN but also for its rehabilitation. While some studies indicate that dual-task training improves gait, cognitive processes, and skill transfer across various neurological disorders [70], others have found no significant additional benefits over single-task training in neglect rehabilitation [71]. The effectiveness of multitasking-based interventions may depend on factors such as training duration, task complexity, and individual patient characteristics. Despite mixed findings, dual-task paradigms remain a promising therapeutic approach in neurological rehabilitation, with the potential to target deficits in both spatial and non-spatial attention [70, 71].

## Materials and Methods

### Participants

The study sample included 102 participants with right hemisphere damage, of whom 38 were diagnosed with visual neglect (VN) and 64 did not exhibit VN. The average age of patients with neglect was 65.6 ± 10.7 years (range: 28–80), and the average age of those without neglect was 60.9 ± 12.3 years (range: 29–82). The sample comprised 46 women and 57 men.

All participants underwent a preliminary comprehensive neuropsychological examination to assess higher mental functions [72], as well as standardized quantitative assessments of visual neglect. Exclusion criteria included severe speech or perceptual disorders, disorientation, confusion, and affective states that interfered with effective interaction with the examiner.

This study was approved by the Research Ethics Committee of the Federal Scientific Center of Psychological and Multidisciplinary Research (Protocol No. 5, May 16, 2024). Prospective recruitment took place between May 17, 2024, and January 30, 2025. Written informed consent was obtained from all participants, including information about voluntary participation, the nature of the study, and permission for the use of anonymized data. Each participant was assigned a unique six-digit code to ensure confidentiality. To prevent the possibility of identifying individuals, one neuropsychologist conducted the assessment, while a second researcher independently received and processed the anonymized data. No minors were included in the sample; all participants were adults recruited in a clinical setting. The study was conducted in accordance with the principles of the Declaration of Helsinki.

### Materials

To carry out the method, a laptop with a screen size of at least 28 cm and a wireless mouse with a mousepad were required. PsychoPy software was used for the study. A study was conducted on a standard laptop with a 15.6-inch screen and Windows 11 system. The method’s functionality was also tested on other operating systems, including Linux (with a recommendation to check the system locale for correct data storage if the system language was English) and Mac OS, as well as on laptops with smaller screen sizes.

### Procedure of One Trial

The main task for the participant is to determine the location of a circle with a diameter of 8 mm. Simultaneously, the participant performs the task of identifying the central digit (1, 2, 3, or 4). Each trial starts with the presentation of a fixation cross for 1000 ms. Then, a circle appears on the periphery, and a digit appears at the center of the screen for 100 ms. Afterward, a black-and-white mask (in the form of a randomly generated dot diagram) appears on the screen for 2000 ms (Fig 1). At the end of one trial, the participant orally reports to the experimenter which digit was in the center of the screen and on which side the circle was located. After the mask, a response input screen appears, where the experimenter enters the participant’s verbal report about what was seen on the screen.

**Figure 1.**
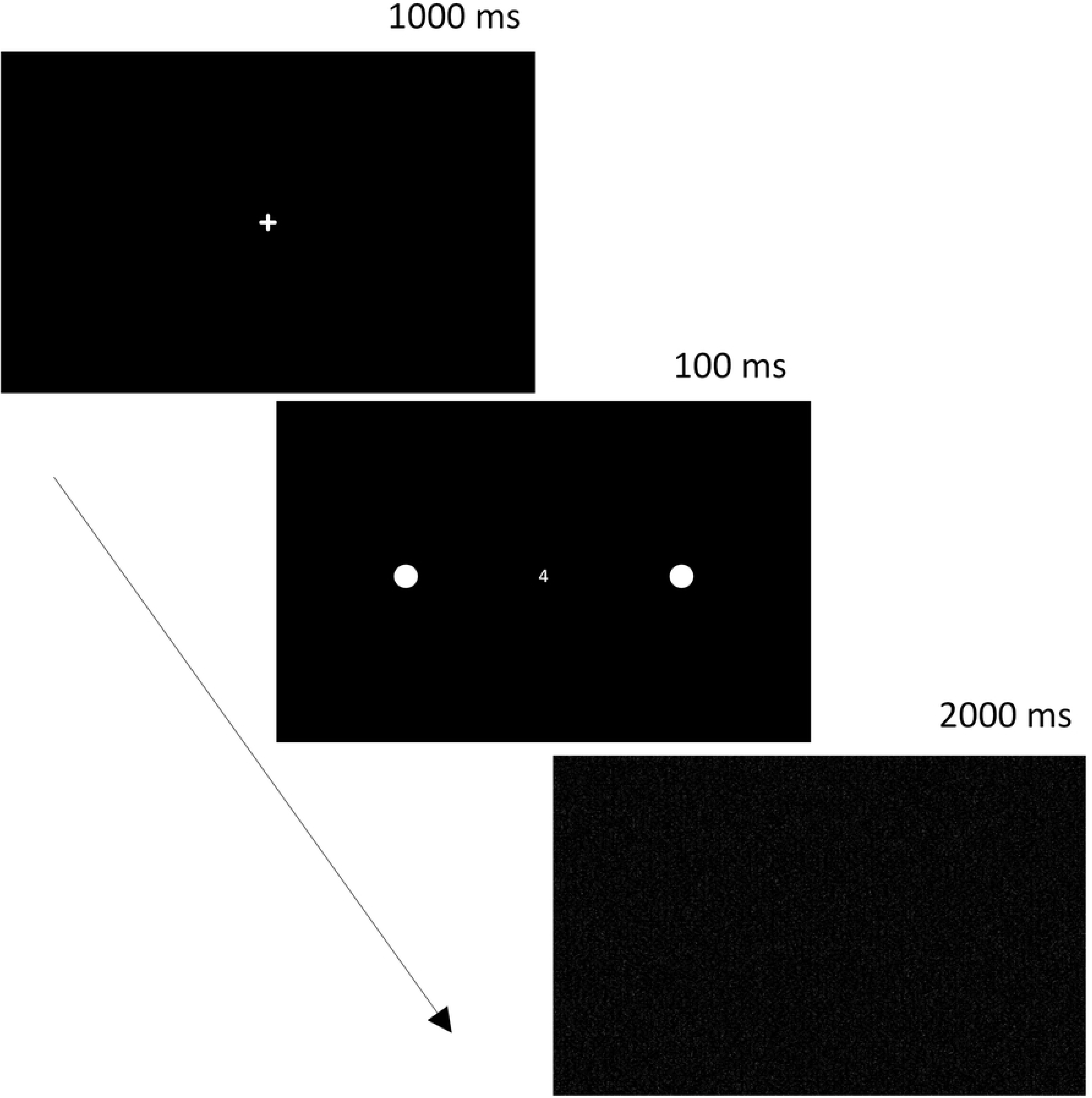
Sequence of events in a single trial.

### Instruction

The participant is given the following standard instruction: “Watch closely what happens on the screen. First, a cross will appear in the center of the screen; focus on it. Then a digit will appear at the cross’s location. Simultaneously with the digit, a circle will appear – either on the right, left, or both sides. The circle can appear in different parts of the screen. Be attentive: the digit and the circle will appear for a very short time! Try to see them. Then say aloud what you managed to notice: the digit and the side where the circle was, for example, ‘One on the right.’”

### Trial Parameters

Before the experiment begins, a screen displaying an example of how the fixation cross, digit, and circle (placed 13.5 cm to the right of the center) will appear is shown. This allows the participants to become familiar with the stimuli and their sizes, and the experimenter can check the settings to ensure proper display of distances on the specific device. At the beginning of the experiment, participants complete a training series consisting of 7 trials. During the training, participants familiarize themselves with the possible trial variations, where parameters such as the number of circles (1 or 2), presentation side, and distance from the screen’s center (both vertically and horizontally) are varied. The stimulus presentation time decreases from 1 second to 100 ms during the training series.

Following the training, the participant proceeds to the main series. The main series consists of 75 trials: 42 unilateral presentations (21 with a circle on the right side of the center and 21 with a circle on the left side) and 33 bilateral presentations. All trials are presented in a randomized order. There are 7 possible positions for the circle on each side, resulting in 14 possible circle positions on the screen (Fig 2).

**Figure 2.**
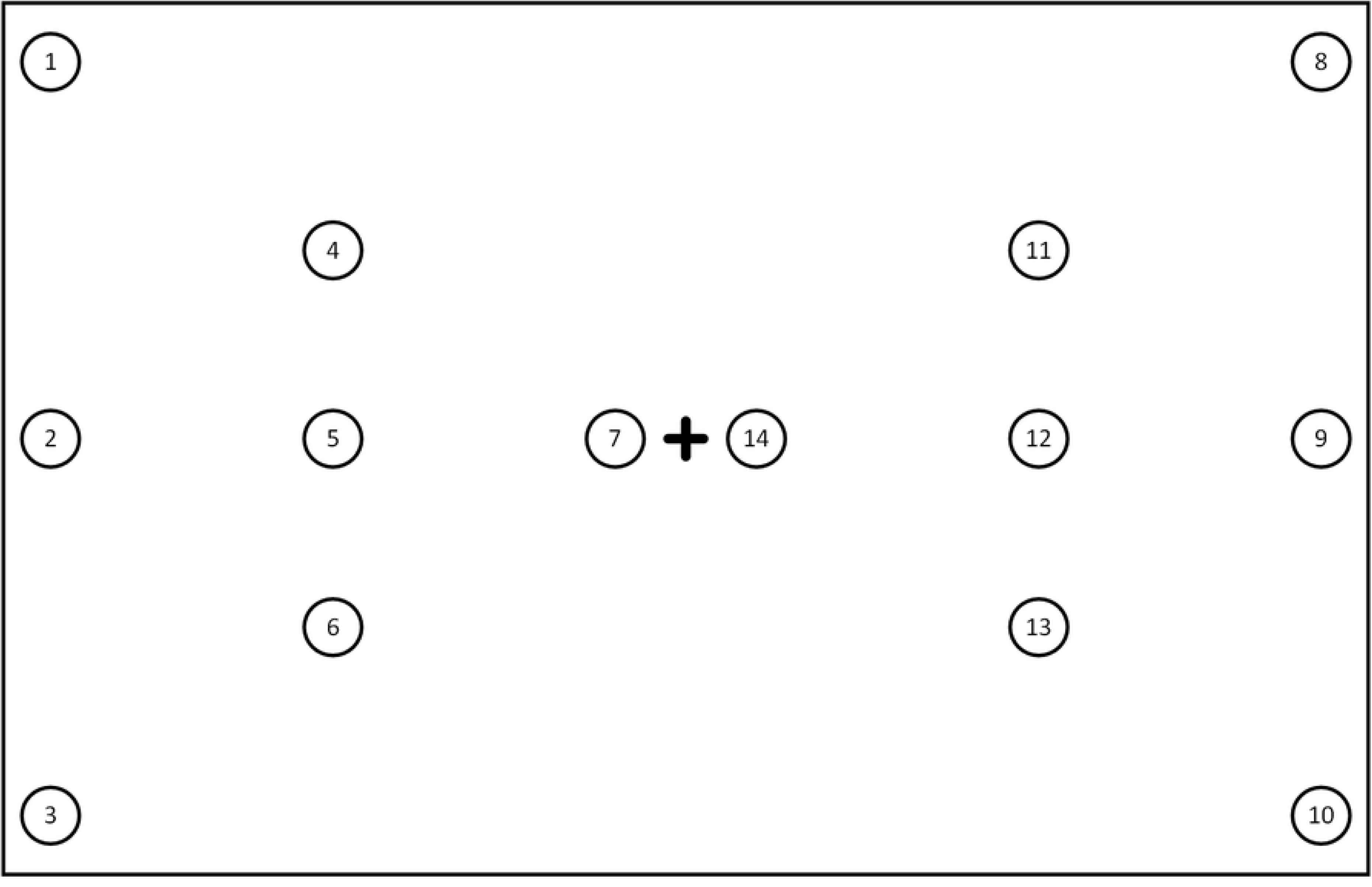
Fixed positions of the target stimulus – circle.

The distance at which each of the 14 circles is located from the center of the screen is presented in Table 1.

**Table 1.** Circle positions relative to the center of the screen.

| Target position number | Horizontal distance from the center of the screen (cm) | Vertical distance from the center of the screen (cm) |
| --- | --- | --- |
| 1 | -13.5 | 8 |
| 2 | -13.5 | 0 |
| 3 | -13.5 | -8 |
| 4 | -7.5 | 4 |
| 5 | -7.5 | 0 |
| 6 | -7.5 | -4 |
| 7 | -1.5 | 0 |
| 8 | 13.5 | 8 |
| 9 | 13.5 | 0 |
| 10 | 13.5 | -8 |
| 11 | 7.5 | 4 |
| 12 | 7.5 | 0 |
| 13 | 7.5 | -4 |
| 14 | 1.5 | 0 |

Table 2 and Fig 3 provide characteristics of all 75 trials in the main series.

**Figure 3.**
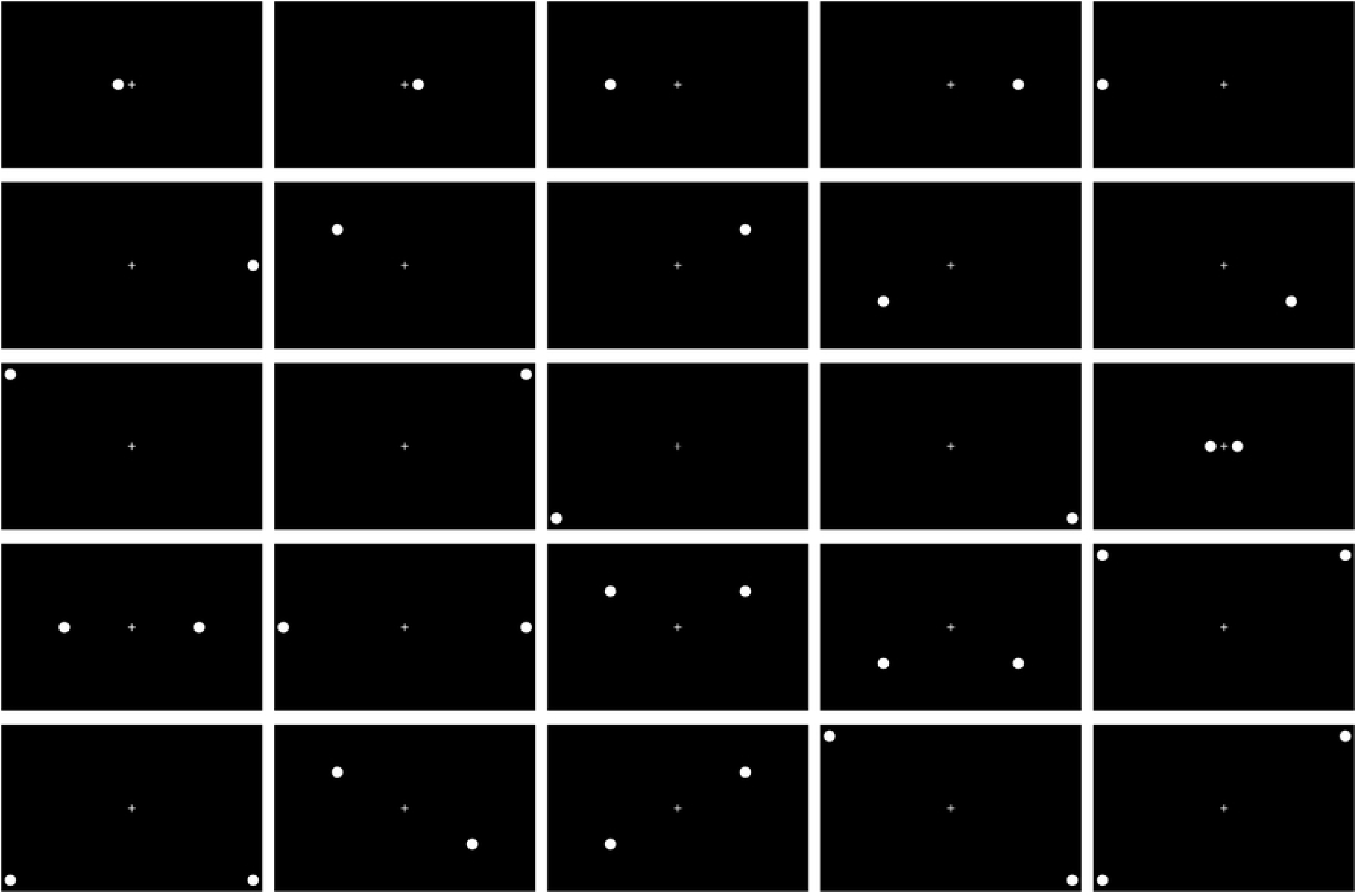
All possible combinations of circles presented in the method.

**Table 2.**
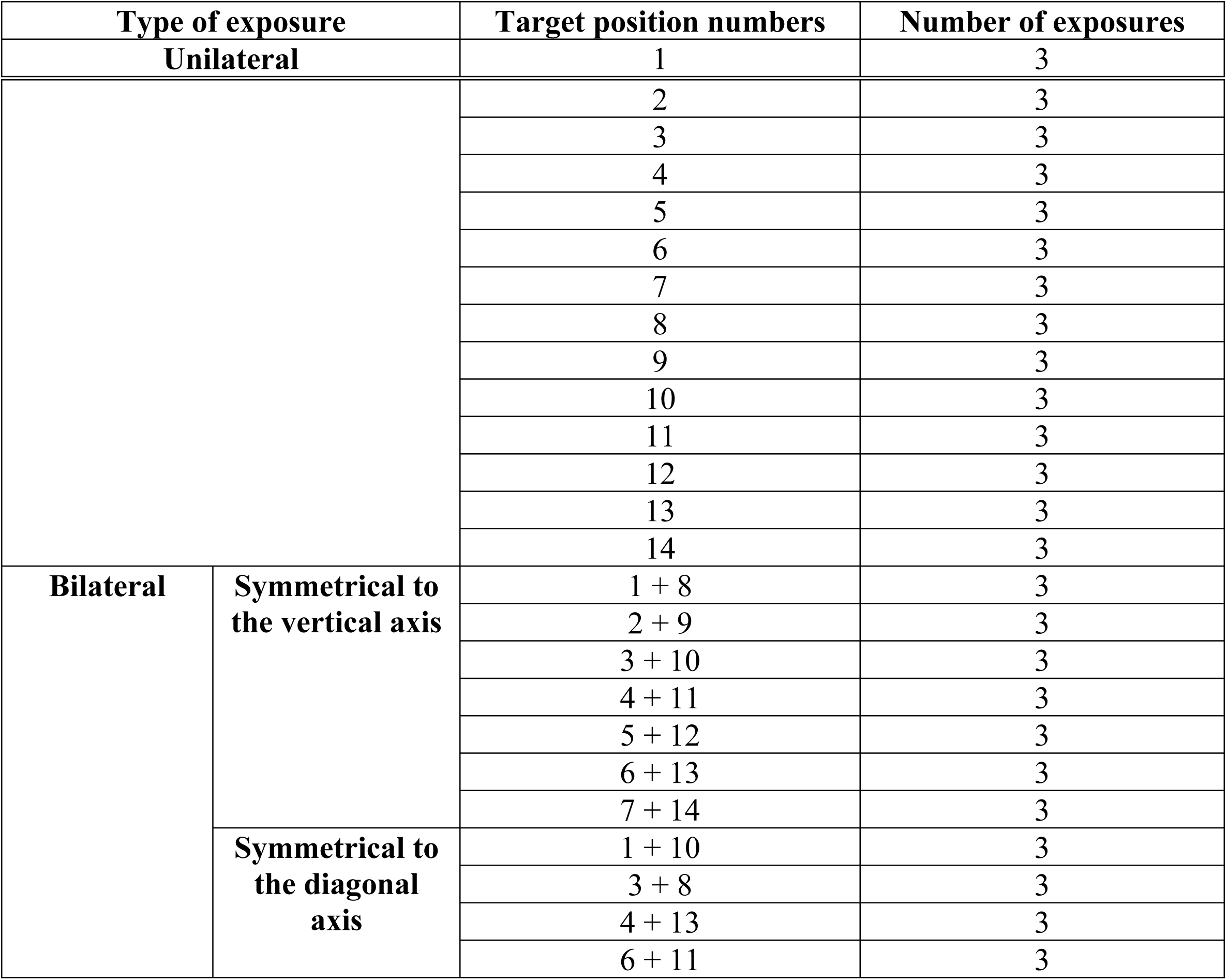
All possible combinations presented in the method.

| Type of exposure | Target position numbers | Number of exposures |
| --- | --- | --- |
| Unilateral | 1 | 3 |

|  |  |  |  |
| --- | --- | --- | --- |
|  |  | 2 | 3 |
|  |  | 3 | 3 |
|  |  | 4 | 3 |
|  |  | 5 | 3 |
|  |  | 6 | 3 |
|  |  | 7 | 3 |
|  |  | 8 | 3 |
|  |  | 9 | 3 |
|  |  | 10 | 3 |
|  |  | 11 | 3 |
|  |  | 12 | 3 |
|  |  | 13 | 3 |
|  |  | 14 | 3 |
| **Bilateral** | **Symmetrical to the vertical axis** | 1 + 8 | 3 |
|  |  | 2 + 9 | 3 |
|  |  | 3 + 10 | 3 |
|  |  | 4 + 11 | 3 |
|  |  | 5 + 12 | 3 |
|  |  | 6 + 13 | 3 |
|  |  | 7 + 14 | 3 |
|  | **Symmetrical to the diagonal axis** | 1 + 10 | 3 |
|  |  | 3 + 8 | 3 |
|  |  | 4 + 13 | 3 |
|  |  | 6 + 11 | 3 |

### Data Processing in the Keen Eye Method

Based on the analysis of the method, the following variables are computed:

- 1-14 – number of errors in the corresponding sectors of the screen (see Fig 2);
- right_skipped_unilateral (RSU) – number of missed right stimuli in the unilateral presentation;
- left_skipped_unilateral (LSU) – number of missed left stimuli in the unilateral presentation;
- right_skipped_bilateral (RSB) – number of missed right stimuli in the bilateral presentation;
- left_skipped_bilateral (LSB) – number of missed left stimuli in the bilateral presentation; right_hallucination (RH) – number of responses indicating “right” when there was no circle on the right side;
- left_hallucination (LH) – number of responses indicating “left” when there was no circle on the left side.

Additionally, similar to dichotic listening [73], the following coefficients are calculated:

- KA_bilateral – asymmetry coefficient in the bilateral presentation;
- KPrR_bilateral – right productivity coefficient in the bilateral presentation;
- KPrL_bilateral – left productivity coefficient in the bilateral presentation;
- KA_all – general asymmetry coefficient;
- KEf_all – general efficiency coefficient;
- KPrR_all – overall right productivity coefficient;
- KPrL_all – overall left productivity coefficient.

The asymmetry coefficients are calculated by the program using the following formulas:

**KA_bilateral** – asymmetry coefficient in the bilateral presentation. This coefficient ranges from −1 to +1 and allows the assessment of the degree of neglect during bilateral stimulus perception. In the absence of a lateralized attention deficit, this parameter should approach 0. It is calculated as:

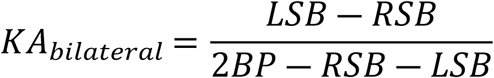

where *BP* equals 33, indicating the number of bilateral presentations.

**KPrR_bilateral** – right productivity coefficient in the bilateral presentation of circles. This describes the proportion of “caught” circles on the right during bilateral presentations and ranges from 0 to 1. It is calculated as:

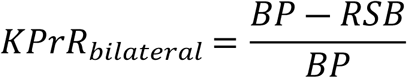

**KPrL_bilateral** – left productivity coefficient in the bilateral presentation of circles. This describes the proportion of “caught” circles on the left during bilateral presentations and ranges from 0 to 1. It is calculated as:

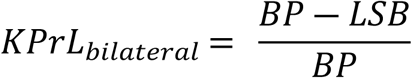

**KA_all** – overall asymmetry coefficient. This coefficient ranges from −1 to +1 and allows the assessment of neglect during both bilateral and unilateral stimulus perceptions. In the absence of a lateralized attention deficit, this parameter should approach 0. It is calculated as:

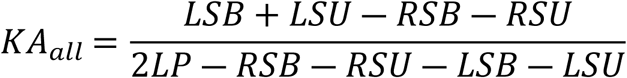

where LP equals 54, indicating the total number of lateralized (left and right) circle presentations.

**KEf_all** – overall efficiency coefficient. It ranges from 0 to 1 and takes into account the number of errors (incorrectly identifying the sides of the presented circles). It is calculated as:

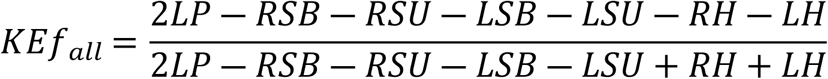

**KPrR_all** – overall right productivity coefficient. This describes the proportion of “caught” circles on the right during all circle presentations and ranges from 0 to 1. It is calculated as:

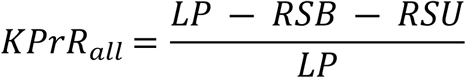

**KPrL_all** – overall left productivity coefficient. This describes the proportion of “caught” circles on the left during all circle presentations and ranges from 0 to 1. It is calculated as:

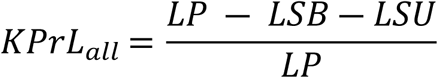

For the justification of convergent validity, Albert’s Test [74] and The Bells Test [29] were selected. These standardized methods were chosen because they are widely recognized techniques used for the quantitative and qualitative diagnosis of patients with unilateral spatial neglect in the visual modality.

### Statistical Analysis

All statistical analyses were performed using RStudio 2023.12.1 Build 402 (packages: rattle, rpart, rpart.plot) and jamovi (version 2.3.18). Descriptive statistics, including mean values, standard deviations, medians, and interquartile ranges, were calculated for all variables. The Mann-Whitney U test was used for non-parametric data. The Kruskal-Wallis test was used for multiple group comparisons, and the post hoc DSCF test with Holm’s correction was applied for multiple comparisons.

The effect size was calculated using rank-biserial correlation for non-parametric tests. To assess the relationship between the severity of neglect and frontal dysfunction, a correlation analysis was performed. Cluster analysis was conducted in jamovi using k-means clustering with silhouette analysis to determine the optimal number of clusters.

## Results

### Comparison Between Groups on the Keen Eye Method Indicators

Descriptive statistics were calculated for all sample parameters. Table 3 presents the mean, standard deviation (SD), minimum, and maximum values for each variable in the control group and the patient group with VN. The results indicate that the average number of omissions on the left side is consistently higher in patients with neglect, with some individuals reaching the maximum possible number of errors on specific measures. Additionally, in patients with VN, the minimum number of errors for certain parameters (e.g., LSB) is notably distant from zero, suggesting variability in performance even among affected individuals.

**Table 3.** Descriptive Statistics for the Control and Visual Neglect Groups.

| Variable | Control Group<br>(M ± SD) | Min - Max | VN group<br>(M ± SD) | Min - Max |
| --- | --- | --- | --- | --- |
| **1** | 2.422 ± 2.580 | 0 - 9 | 6.289 ± 1.228 | 3 - 9 |
| **2** | 1.281 ± 1.598 | 0 - 6 | 3.421 ± 1.154 | 1 - 6 |
| **3** | 2.594 ± 2.975 | 0 - 9 | 6.263 ± 1.519 | 2 - 9 |
| **4** | 1.609 ± 2.201 | 0 - 7 | 5.842 ± 1.443 | 2 - 9 |
| **5** | 0.968 ± 1.333 | 0 - 4 | 3.211 ± 1.212 | 0 - 6 |
| **6** | 2.016 ± 2.427 | 0 - 9 | 6.000 ± 1.644 | 2 - 9 |
| **7** | 0.688 ± 1.022 | 0 - 3 | 3.184 ± 1.557 | 0 - 6 |
| **8** | 2.922 ± 3.184 | 0 - 9 | 7.974 ± 1.684 | 3 - 9 |
| **9** | 1.766 ± 2.114 | 0 - 6 | 5.342 ± 1.097 | 2 - 6 |
| **10** | 2.969 ± 3.266 | 0 - 9 | 8.289 ± 1.137 | 6 - 9 |
| **11** | 2.266 ± 2.857 | 0 - 9 | 7.421 ± 2.176 | 1 - 9 |
| **12** | 1.359 ± 1.947 | 0 - 6 | 4.868 ± 1.436 | 1 - 6 |
| **13** | 2.141 ± 2.689 | 0 - 9 | 7.974 ± 1.636 | 2 - 9 |
| **14** | 0.953 ± 1.430 | 0 - 6 | 4.395 ± 1.994 | 0 - 6 |
| **RSU** | 1.922 ± 2.978 | 0 - 14 | 4.289 ± 4.299 | 0 - 16 |
| **LSU** | 4.656 ± 5.782 | 0 - 21 | 16.368 ± 5.749 | 1 - 21 |
| **RSB** | 4.531 ± 6.510 | 0 - 32 | 6.500 ± 6.479 | 0 - 27 |
| **LSB** | 7.953 ± 10.045 | 0 - 33 | 29.815 ± 4.980 | 14 - 33 |
| **NIR** | 3.984 ± 4.971 | 0 - 21 | 10.500 ± 8.013 | 0 - 21 |
| **NIL** | 3.609 ± 4.699 | 0 - 21 | 9.816 ± 7.651 | 0 - 21 |
| **NIB** | 5.250 ± 6.785 | 0 - 33 | 12.421 ± 11.337 | 0 - 33 |
| **RH** | 0.391 ± 0.866 | 0 - 5 | 1.632 ± 3.062 | 0 - 16 |
| **LH** | 0.125 ± 0.333 | 0 - 1 | 0.132 ± 0.343 | 0 - 1 |
| **KA_bilateral** | 0.101 ± 0.324 | -0.941 - 1 | 0.826 ± 0.239 | 0.136 - 1 |
| **KPrR_bilateral** | 0.863 ± 0.197 | 0.030 - 1 | 0.803 ± 0.196 | 0.182 - 1 |
| **KPrL_bilateral** | 0.759 ± 0.304 | 0 - 1 | 0.097 ± 0.151 | 0 - 0.576 |
| **KA_all** | 0.104 ± 0.274 | -0.479 - 0.927 | 0.746 ± 0.247 | 0.182 - 1 |
| **KEf_all** | 0.987 ± 0.033 | 0.760 - 1 | 0.927 ± 0.126 | 0.407 - 1 |
| **KPrR_all** | 0.881 ± 0.164 | 0.278 - 1 | 0.800 ± 0.197 | 0.204 - 1 |
| **KPrL_all** | 0.767 ± 0.285 | 0.037 - 1 | 0.145 ± 0.172 | 0 - 0.667 |

A comparison was made between groups of right hemisphere patients without neglect and those with neglect using the Mann-Whitney U test. The results of the analysis are presented in Table 4. Significant differences, accounting for Holm’s correction, were found in all indicators (p<0.01), except for LH (p=0.684) and RSB (p=0.096). Among the error omission sums, the most important indicators are LSB and LSU, which highlights the specificity of this task for patients with left-sided neglect. Both indicators are important, but they can dissociate, allowing for the diagnosis not only of neglect but also of visual extinction.

**Table 4.**
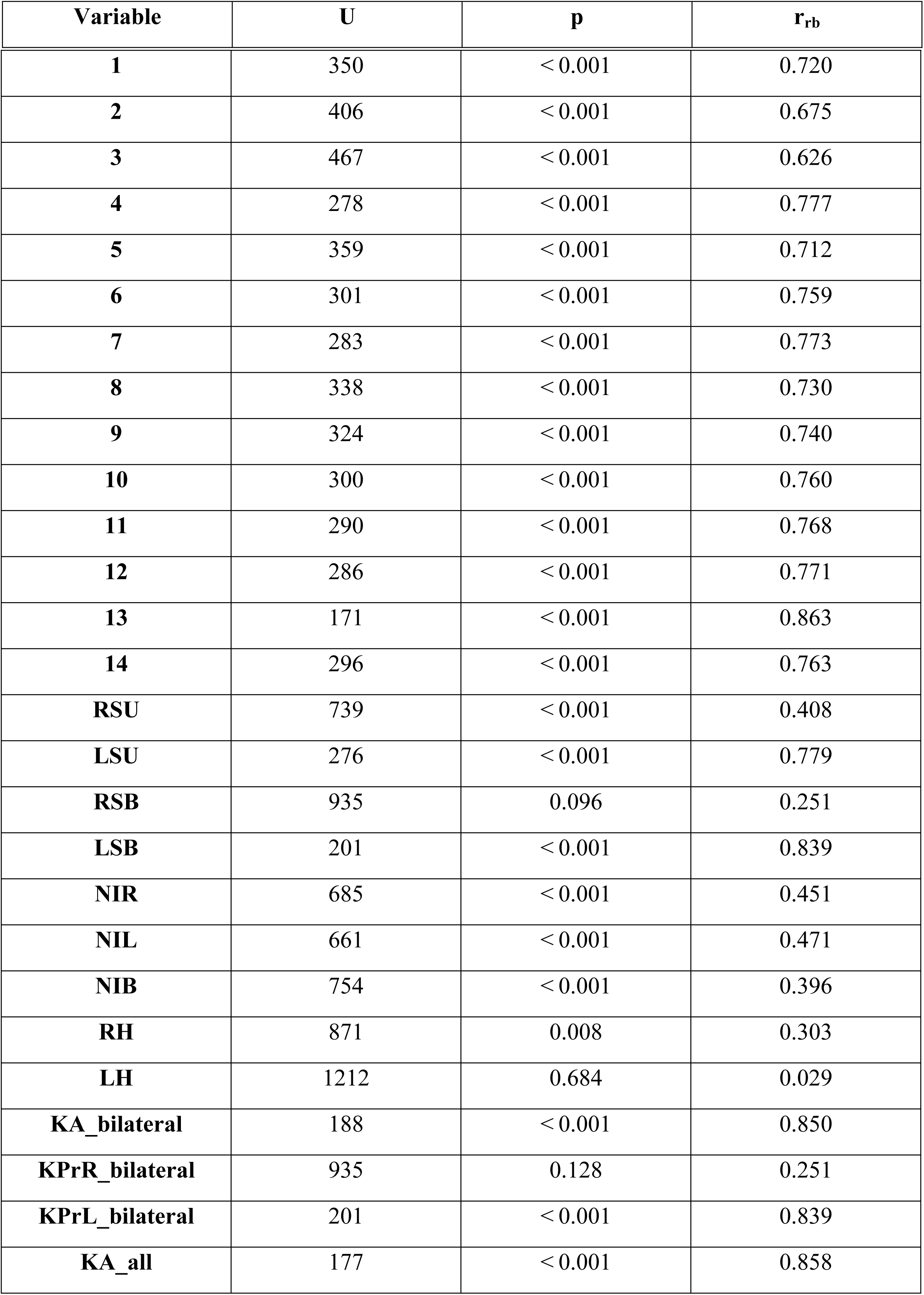
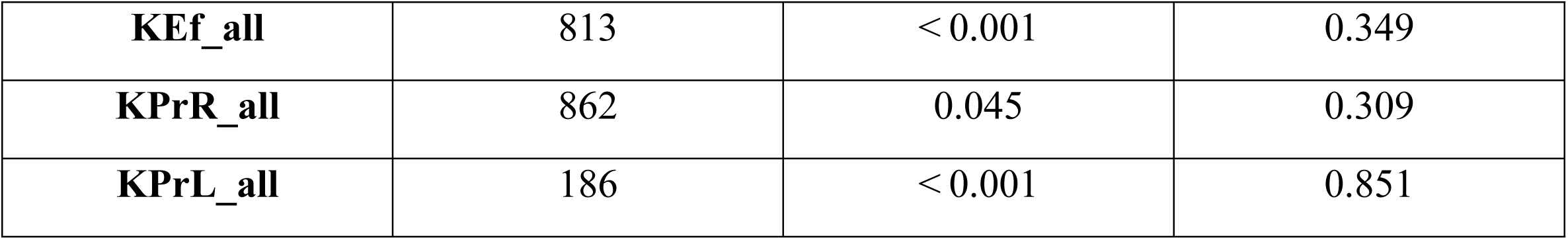
Intergroup Comparisons of All Parameters Using the Mann-Whitney Test and Rank-Biserial Correlation.

| Variable | U | p | r<sub>rb</sub> |
| --- | --- | --- | --- |
| **1** | 350 | < 0.001 | 0.720 |
| **2** | 406 | < 0.001 | 0.675 |
| **3** | 467 | < 0.001 | 0.626 |
| **4** | 278 | < 0.001 | 0.777 |
| **5** | 359 | < 0.001 | 0.712 |
| **6** | 301 | < 0.001 | 0.759 |
| **7** | 283 | < 0.001 | 0.773 |
| **8** | 338 | < 0.001 | 0.730 |
| **9** | 324 | < 0.001 | 0.740 |
| **10** | 300 | < 0.001 | 0.760 |
| **11** | 290 | < 0.001 | 0.768 |
| **12** | 286 | < 0.001 | 0.771 |
| **13** | 171 | < 0.001 | 0.863 |
| **14** | 296 | < 0.001 | 0.763 |
| **RSU** | 739 | < 0.001 | 0.408 |
| **LSU** | 276 | < 0.001 | 0.779 |
| **RSB** | 935 | 0.096 | 0.251 |
| **LSB** | 201 | < 0.001 | 0.839 |
| **NIR** | 685 | < 0.001 | 0.451 |
| **NIL** | 661 | < 0.001 | 0.471 |
| **NIB** | 754 | < 0.001 | 0.396 |
| **RH** | 871 | 0.008 | 0.303 |
| **LH** | 1212 | 0.684 | 0.029 |
| **KA_bilateral** | 188 | < 0.001 | 0.850 |
| **KPrR_bilateral** | 935 | 0.128 | 0.251 |
| **KPrL_bilateral** | 201 | < 0.001 | 0.839 |
| **KA_all** | 177 | < 0.001 | 0.858 |
| **KEf_all** | 813 | < 0.001 | 0.349 |
| **KPrR_all** | 862 | 0.045 | 0.309 |
| **KPrL_all** | 186 | < 0.001 | 0.851 |

Table 4 presents the results of the intergroup comparison for the asymmetry coefficients. Among the asymmetry coefficients, KA_all and KA_bilateral are the most significant and may be used as indicators of neglect severity. In contrast to the omission error count, this indicator is independent of the number of stimuli and serves as a convenient ordinal index of neglect severity, ranging from −1 to 1.

### Diagnostic Criterion Selection Using Decision Trees

To identify the most significant diagnostic criteria for neglect using the Keen Eye method, decision tree analyses were conducted. This approach allows data classification based on features selected from the original dataset, with the goal of building a predictive model for diagnosis using specified predictor variables. The following parameters were used to evaluate model quality: accuracy, precision, recall, and F1 score.

The predictors for classification models 1 and 2 included total error counts (RSU, LSU, RSB, LSB, RH, and LH). Model 1 (Fig 4) was constructed using cross-validation (Accuracy = 0.90, Precision = 0.82, Recall = 0.95, F1 Score = 0.88, Sensitivity = 0.95, Specificity = 0.88).

**Figure 4.**
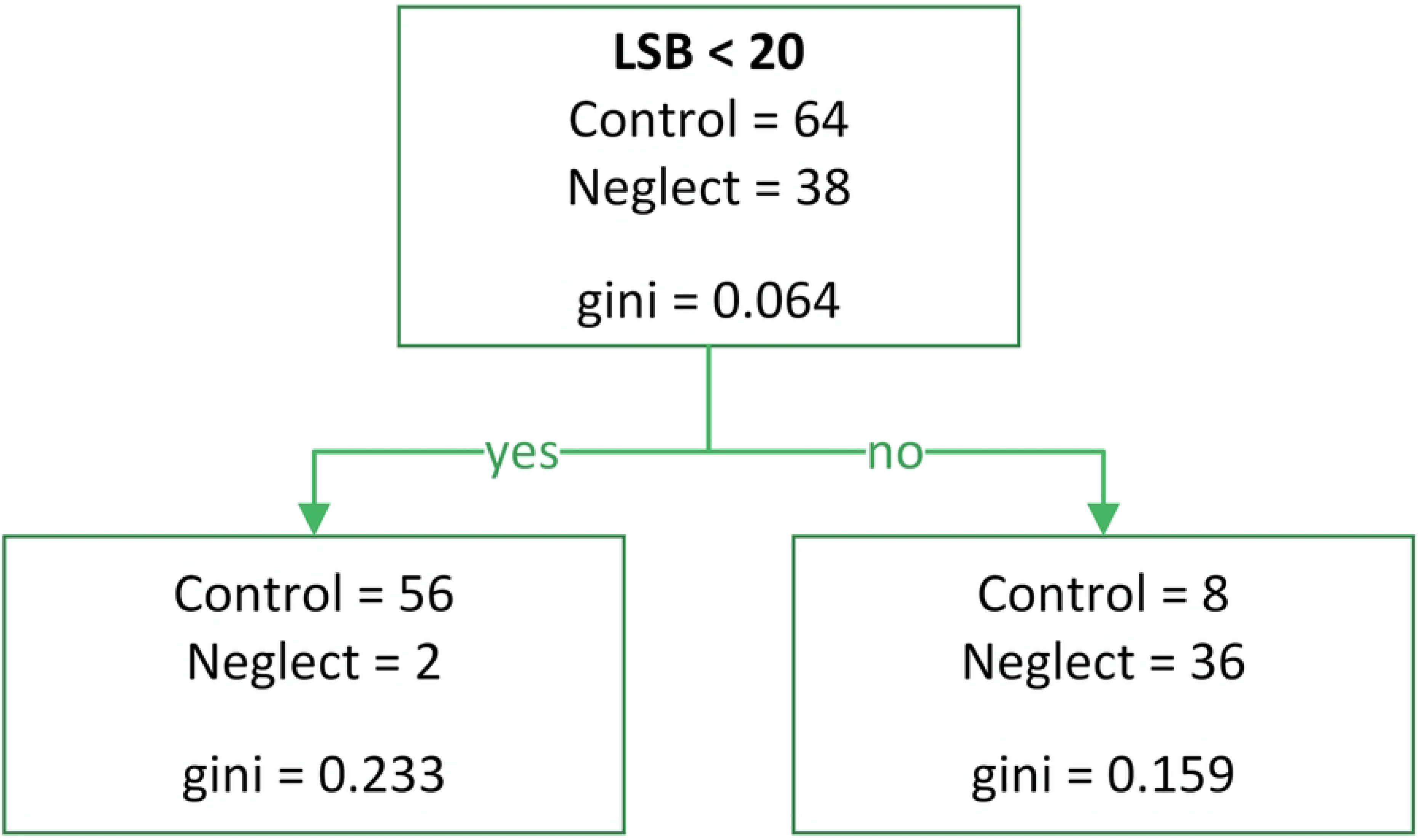
Decision Tree Model 1 for Predicting Visual Neglect Diagnosis.

Model 2 (Fig 5) was designed to maximize recall and improve specificity (Accuracy = 0.87, Precision = 0.75, Recall = 1.00, F1 Score = 0.85, Sensitivity = 1.00, Specificity = 0.80).

**Figure 5.**
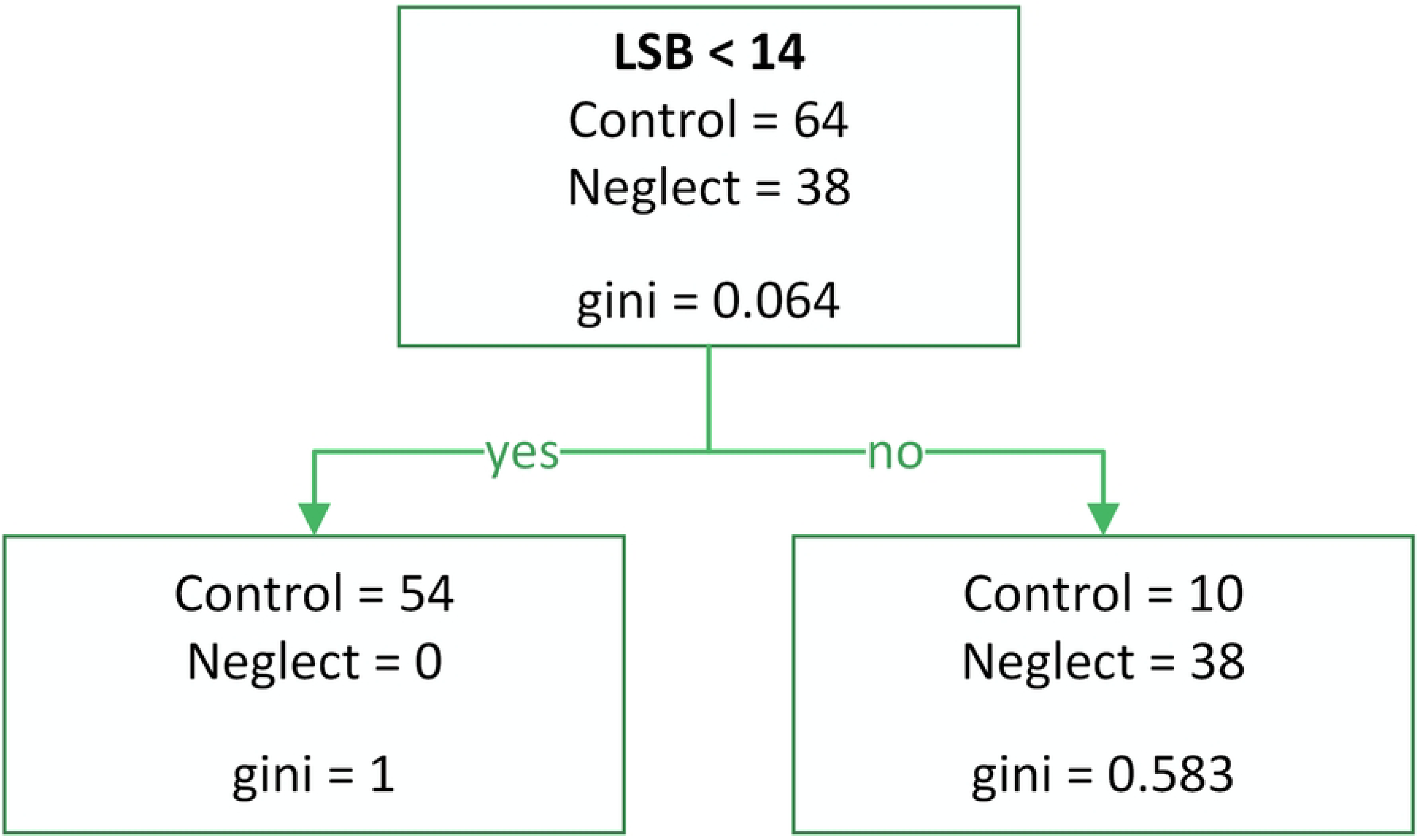
Decision Tree Model 2 for Predicting Visual Neglect Diagnosis.

Model 3 (Fig 6) was developed using predictors based on asymmetry coefficients (KA_bilateral, KPrR_bilateral, KPrL_bilateral, KA_all, KEf_all, KPrR_all, and KPrL_all) (Accuracy = 0.90, Precision = 0.79, Recall = 1.00, F1 Score = 0.88, Sensitivity = 1.00, Specificity = 0.84).

**Figure 6.**
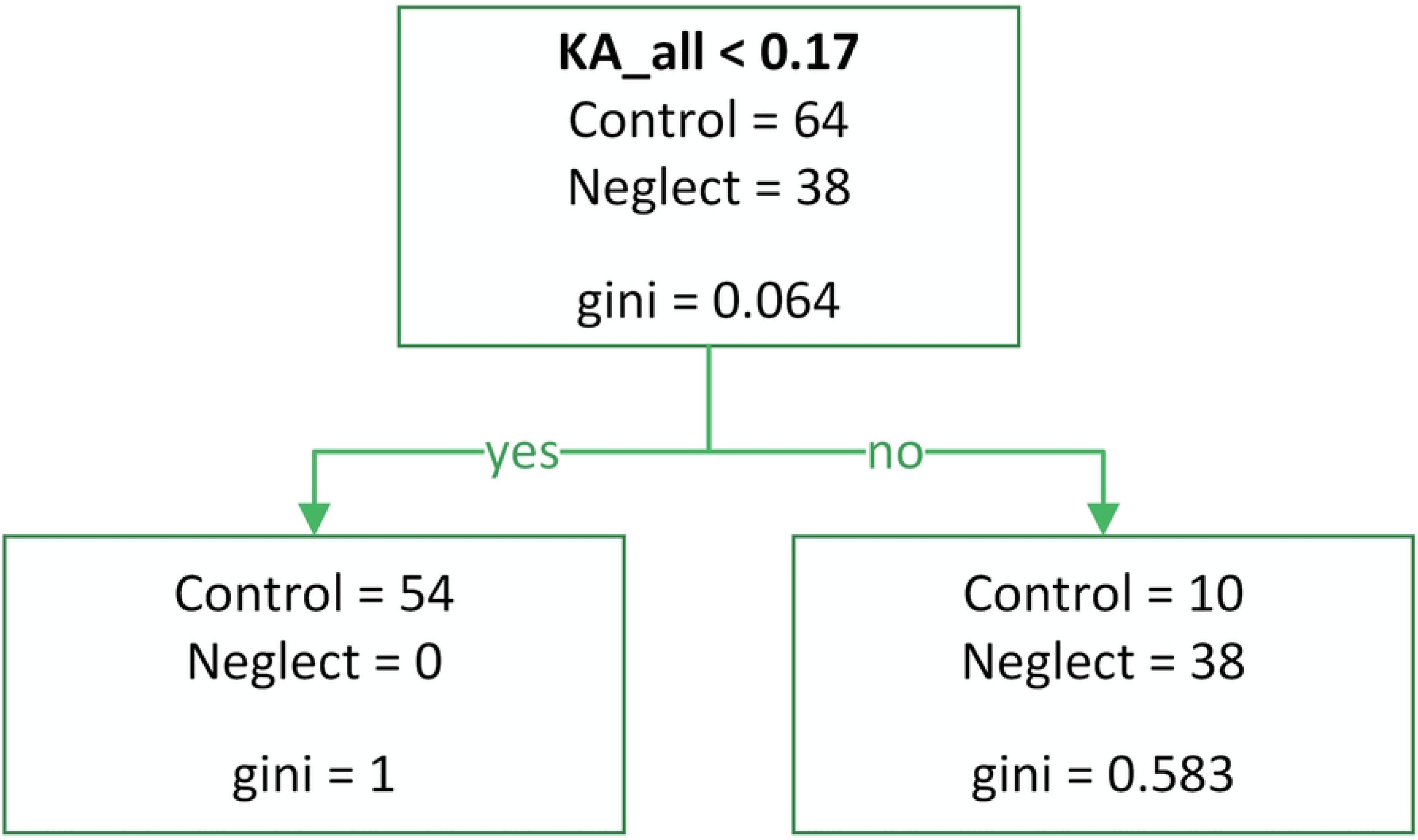
Decision Tree Model 3 for Predicting Visual Neglect Diagnosis.

All three models demonstrated high sensitivity; however, some patients without a diagnosed neglect condition were incorrectly classified into the neglect group. This may suggest that the Keen Eye method is capable of detecting hidden forms of VN, including well-compensated neglect.

From a diagnostic standpoint, the most important indicator is LSB. The LSB value is particularly useful for identifying visual extinction and can dissociate from LSU when a patient recognizes a single left-side presentation but fails to acknowledge the left side when two objects are presented in the perceptual field. When using Models 2 and 3 for diagnosing neglect, the method exhibits high specificity.

### Diagnosis of Vertical Neglect Using the Keen Eye Method

To identify vertical neglect, we selected 48 patients who, according to the Keen Eye method, were classified as “neglect” based on the criterion KA_all ≥ 0.17 (Model 3).

The following indicators were calculated to detect vertical neglect:

- up: the number of missed circles in positions above the horizontal line (positions 1, 4, 8, 11).
- down: the number of missed circles in positions below the horizontal line (positions 3, 6, 10, 13).

According to the literature, the left lower portion of the visual field tends to be neglected. Therefore, we used a directional hypothesis (down > up). The Wilcoxon signed-rank test was used to compare the related samples.

Results from the within-group comparison showed significant differences (W = 251, df = 47, p = 0.025). The mean number of left-sided lower and upper omissions in the group of patients with neglect is presented in Fig 7.

**Figure 7.**
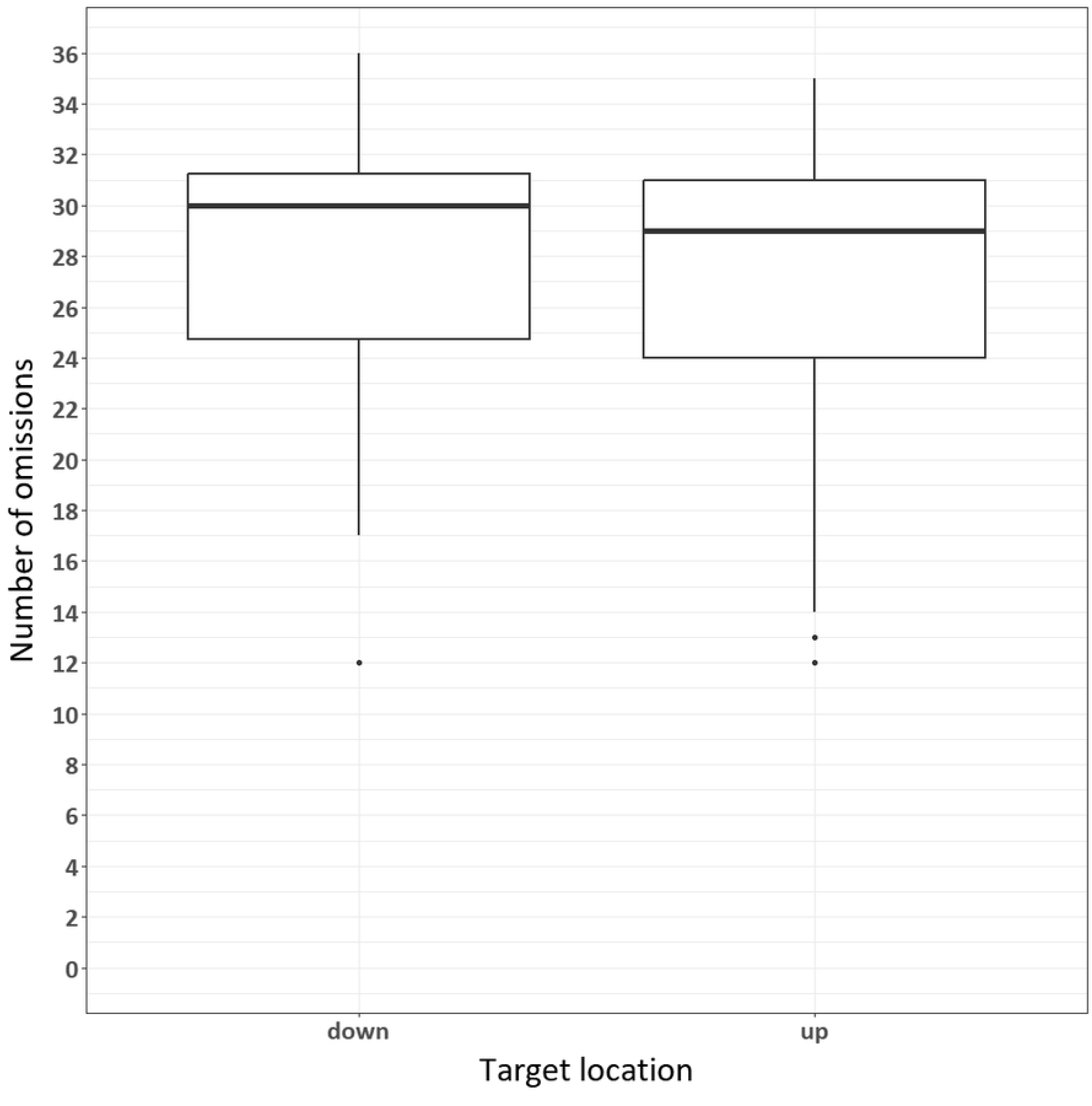
Descriptive graph of vertical misses (up vs. down) in the neglect group.

Thus, the Keen Eye method aligns with findings of predominant neglect in the lower-left visual field in patients with neglect.

### Criterion and Construct Convergent Validity

Convergent validity was assessed using Guilford’s Phi coefficient. The calculation was based on a comparison between the neuropsychologist’s diagnosis and the diagnosis derived from test performance. The Keen Eye method identified LSB ≥ 12 as the threshold for diagnosing neglect, as determined by the classification tree method. Neglect was also diagnosed based on performance in paper-and-pencil tests, including the Bells Test and Albert’s Test. Table 5 presents the correlation between the Keen Eye results and traditional neglect assessments.

**Table 5.** Correlation Between Keen Eye Results and Paper-and-Pencil Tests (Bells Test and Albert’s Test) Using the Phi Coefficient.

|  | **diagnosis** | **Model 1** | **Model 2** | **Model 3** | **Albert's Test** | **The Bell's Test** |
| --- | --- | --- | --- | --- | --- | --- |
| **Diagnosis** |  |  |  |  |  |  |
| **Model 1** | 0.802*** |  |  |  |  |  |
| **Model 2** | 0.771*** | 0.871*** |  |  |  |  |
| **Model 3** | 0.817*** | 0.924 *** | 0.943 *** |  |  |  |
| **Albert's Test** | 0.481*** | 0.487*** | 0.512*** | 0.513*** |  |  |
| **The Bell's Test** | 0.515*** | 0.496*** | 0.469*** | 0.473*** | 0.570*** |  |

Note. \* $p < 0.05$ , \*\* $p < 0.01$ , \*\*\* $p < 0.001$

**Table 6.** Correlation Between Asymmetry Coefficients, Digit Omissions, and FAB Scores Using Spearman’s Rho.

|  | FAB | KA_bilateral | KA_all | NIR | NIL | NIB |
| --- | --- | --- | --- | --- | --- | --- |
| FAB |  |  |  |  |  |  |
| KA_bilateral | 0.384*** |  |  |  |  |  |
| KA_all | 0.363*** | 0.960*** |  |  |  |  |
| NIR | 0.436*** | 0.401*** | 0.434*** |  |  |  |
| NIL | 0.339*** | 0.388*** | 0.422*** | 0.850*** |  |  |
| NIB | -0.327*** | 0.379*** | 0.402*** | 0.742*** | 0.690*** |  |

Note. \* $p < 0.05$ , \*\* $p < 0.01$ , \*\*\* $p < 0.001$

From the perspective of criterion validity, the Keen Eye method shows high scores, surpassing the diagnostic results of The Bell’s Test and Albert’s Test, which also suggests high incremental and concurrent validity for the computer-based method. From the perspective of convergent validity, the Keen Eye test shows moderate similarity to The Bell’s Test and Albert’s Test. This is explained by the fact that the paper-and-pencil tests are designed to diagnose a more severe form of left-sided neglect, and many patients perform these tests with few or no errors. During paper-and-pencil testing, patients may use compensatory strategies, and the time to complete the tasks is not limited. In contrast, when using the computer-based method, it is virtually impossible to use compensatory strategies for visual search due to the time constraints imposed on stimulus presentation.

### Cluster Analysis of Patients Based on Keen Eye Results

A k-means cluster analysis was conducted using the parameters RSU, LSU, RSB, LSB, NIR, NIL, NIB, RH, and LH. The analysis identified three distinct patient groups (Fig 8):

1. Patients with minimal errors, demonstrating near-perfect accuracy in both circle and number identification.
2. Patients with a high number of circle omissions on the left but accurate number identification.
3. Patients with a high number of circle omissions on the left, in both unilateral and bilateral presentations, along with errors in number identification.

**Figure 8.**
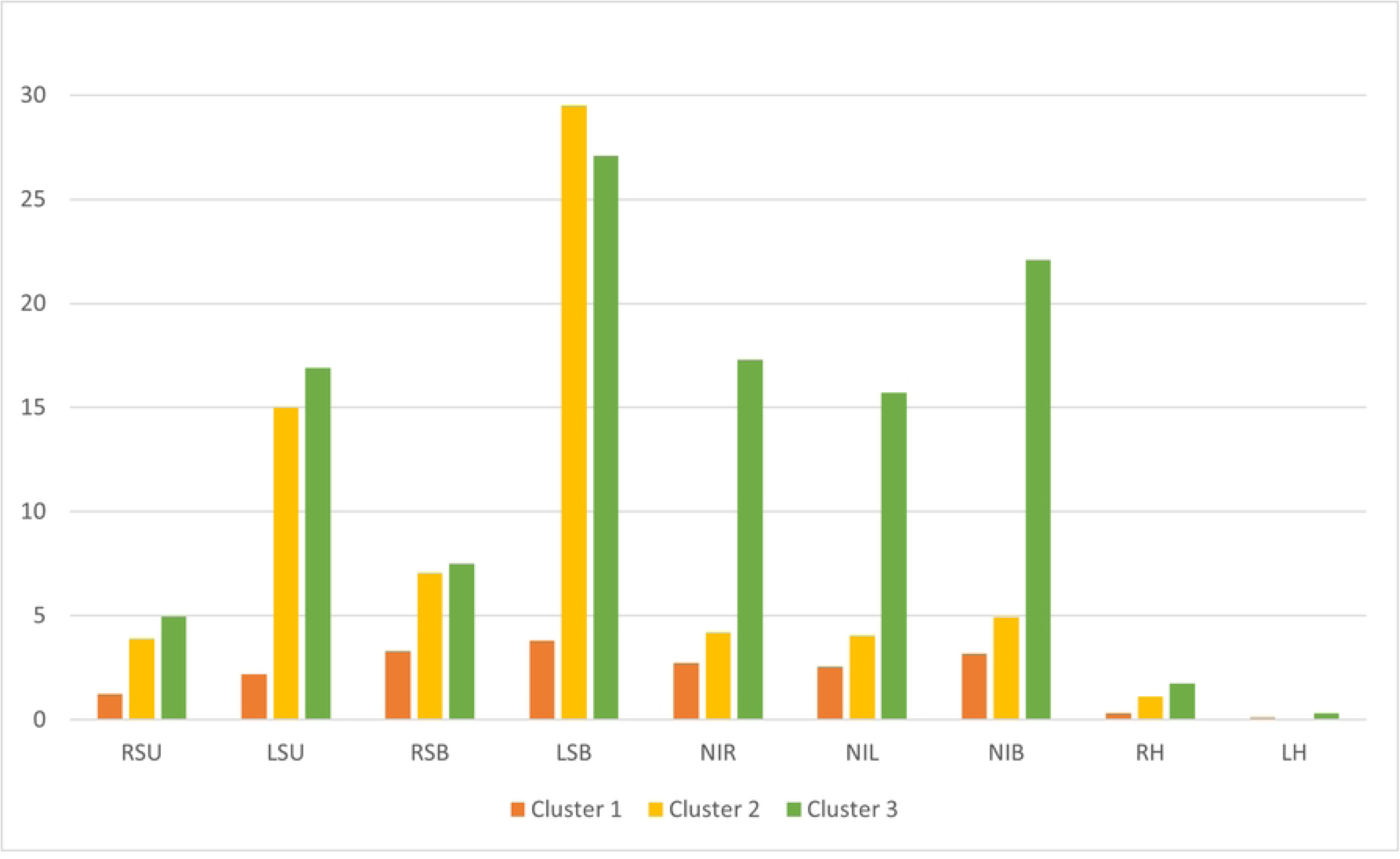
Mean Values of Keen Eye Parameters Across Clusters 1, 2, and 3.

Thus, the Keen Eye method allows for the identification not only of a lateralized attention deficit but also of a general reduction in cognitive resources for attention allocation, which, according to studies, is also characteristic of patients with neglect [58, 75].

### Relationship Between Attention Distribution Deficit Across Two Tasks and Frontal Dysfunction

The identification of Cluster 3 (patients with difficulties in distributing attention across two tasks) raised the question of whether these types of errors are related to impairments in executive functions. We hypothesized that omissions or incorrect identification of digits may be associated with frontal dysfunction. However, omissions in digit identification could also result from a non-lateralized attention deficit, which is theorized to be linked to neglect severity. To test this hypothesis, we conducted a correlation analysis between FAB battery scores [76], neglect severity indicators assessed using the Keen Eye method (KA_bilateral, KA_all), and errors in digit identification (NIR, NIL, NIB).

The correlation analysis revealed a moderate relationship between digit identification errors and frontal dysfunction, as well as a moderate relationship between digit identification errors and the severity of neglect.

Subsequently, a non-parametric Kruskal-Wallis test was conducted to determine whether Cluster 1 differed from Clusters 2 and 3 in terms of FAB scores. Significant results were obtained using the Kruskal-Wallis test (χ2 = 18.1, p < 0.001, η² = 0.19). Post hoc pairwise comparisons using the DSCF test with Holm’s correction revealed significant differences between Cluster 1 and Cluster 2 (W = −5.029, p = 0.03), and between Cluster 1 and Cluster 3 (W = −5.017, p = 0.02). No significant differences were found between Clusters 2 and 3. The mean number of FAB scores obtained by patients from different clusters is presented in Fig 9.

**Figure 9.**
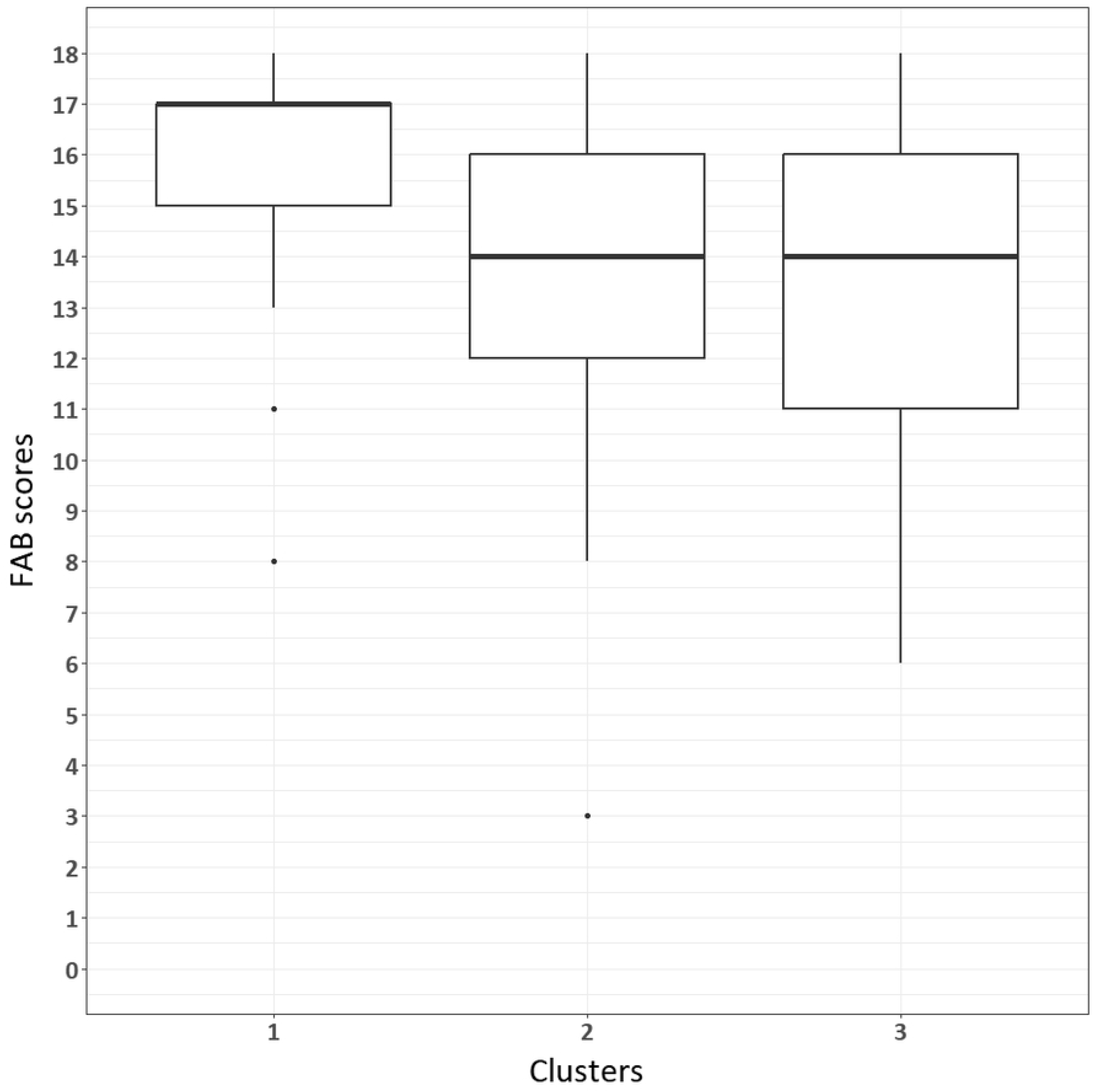
FAB scores between clusters.

These pairwise comparisons demonstrate that both neglect patient clusters (Clusters 2 and 3) have significantly lower FAB scores compared to Cluster 1, but no differences were found between the two neglect clusters.

## Discussion

The results obtained indicate that the Keen Eye method is suitable for detecting VN. The most diagnostically significant indicators are the omissions of circles to the left of the center of the screen during unilateral and bilateral presentations. These omissions allow for a quantitative assessment not only of inattention to single stimuli on the left but also of visual extinction phenomena. Additionally, the method allows for the calculation of various asymmetry coefficients for each subject. It has been shown that the most crucial criterion for diagnosing neglect is the KA_all coefficient. This coefficient is convenient for quantitatively expressing the severity of VN, as it varies from −1 to 1, with a value of 0 for healthy subjects.

Thanks to the various positions of the circles relative to the horizontal line, the method can detect vertical neglect. It has been shown that patients with VN predominantly ignore the lower-left portion of the space. This finding aligns with literature data, according to which vertical neglect is often combined with horizontal neglect and primarily affects the lower part of the visual field [16, 18]. This phenomenon is linked to both cortical and subcortical lesions, especially in the temporal lobe [16]. Vertical neglect can manifest as an allocentric form of spatial neglect: it is proposed that allocentric representations influence the distribution of attention resources in the vertical dimension [77]. Patients with VN demonstrate impairment in automatic, externally controlled covert attention orientation in the contralesional visual field, particularly in the lower quadrant [18]. This vertical shift, it seems, depends on time: patients require more visual fixations and more time to find a target in the lower-left quadrant [23]. This suggests that primarily involuntary automatic attention is impaired.

The Keen Eye method demonstrates high sensitivity compared to existing paper-and-pencil methods that are predominantly used for neuropsychological diagnosis of VN. The Keen Eye method uncovers hidden forms of VN that are not detected during paper-and-pencil testing. Notably, it is important to emphasize that Keen Eye reduces the influence of compensatory strategies due to strict time limitations: with stimuli presented for 100 ms, patients cannot perform voluntary visual search for the target. The high sensitivity of the method is ensured by the increased load on the attention distribution system through the simultaneous presentation of two tasks. Studies show that multitasking and increased cognitive load can reveal hidden forms of spatial neglect in patients who have had a stroke. It has been shown that a dual-task paradigm, combining spatial monitoring with additional tasks, enhances the contralesional deficit in patients with damage to either hemisphere, even when standard tests show no impairments [69, 79]. Such computer-based methods can uncover subclinical VN and extinction along both the horizontal and vertical axes [64, 69]. The exacerbation of VN under increased cognitive load may be associated with deficits in spatial and sustained attention following damage to the right hemisphere [78].

Multitasking and high cognitive load lead to disruptions in attention processes even in healthy individuals [41, 42], but these effects are more pronounced in patients with right hemisphere lesions [55, 56]. It has been shown that the right hemisphere plays a crucial role in multitasking, and its damage results in a chronic deficit in distributing attention across multiple tasks, even after the regression of other symptoms [58]. Thus, limitations in multitasking are natural for a healthy brain, but in pathologies such as stroke, cognitive resources are significantly more impaired. Cognitive reserve, which typically protects against the consequences of brain damage, becomes less effective under multitasking conditions, making it possible to identify the “borderline area” between hidden and overt behavioral impairments [62]. This is especially important for diagnosing subclinical forms of VN, which can only be detected under increased cognitive load.

Moreover, the RH and LH parameters allow for the diagnosis of optical allesthesia, which is impossible in paper-and-pencil diagnostics. Optical allesthesia, or allochiria [80], is a rare phenomenon in which visual stimuli are mislocalized on the opposite side and can occur in patients with neglect. Studies have shown that increased attention load can cause false spatial localization, including allesthesia, in patients with right hemisphere damage [81]. Allochiria in VN can manifest in various modalities [82], which can be explained by a disturbance in mental spatial representation [83]. Another potential explanation is the model of attention disturbance in neglect syndrome, according to which the body-centered matrix responsible for controlling spatial attention is impaired [84]. Altered attention distribution in neglect occurs due to changes in the neural representation of egocentric space. Normally, any action in space, including voluntary control of attention via eye, head, and hand movements, is organized based on the body’s position. To acquire information about the environment in an egocentric coordinate system, integration of sensory information from various spatial sources is required. The primary afferent signals related to the egocentric coordinate system are proprioceptive signals from the neck muscles and vestibular information. In patients with neglect, the process of information integration occurs with systematic errors, leading to a rotation of the entire egocentric reference system around the vertical axis to a new “equilibrium position” on the right side. It is suggested that neglect results from damage to cortical structures responsible for converting this sensory information into egocentric internal “reference matrices.” It has been shown that vestibular stimulation reduces neglect symptoms [85].

Additionally, the method does not require motor responses from the subjects, allowing it to diagnose patients with motor impairments and differentiate between motor and visual forms of neglect. Research shows that with traditional paper-and-pencil tests, it is difficult to distinguish between motor and sensory neglect. It was found that neglect is typically associated with the “input channel,” but the temporal and spatial aspects of neglect related to the “output channel” may require more nuanced differentiation [85]. Research also demonstrates that task-specific requirements influence the classification of attention subtypes [86]. Additionally, researchers have described differences in attention deficits between groups with perceptual and motor neglect, emphasizing the need to use both paper-and-pencil and computer-based tests to identify spatially specific and non-lateralized attention deficits [35]. Collectively, these studies highlight the complexity of assessing neglect and the limitations of traditional tests, underscoring the need for more comprehensive and sensitive assessment methods to accurately differentiate motor and sensory neglect subtypes.

Cluster analysis of patients based on stimulus omission rates and identification errors revealed three distinct groups. One group included patients with a non-lateralized attention deficit, characterized by difficulties in distributing attention between two tasks. Another group demonstrated classic signs of VN without pronounced difficulties in attention distribution. Interestingly, patients with a non-lateralized attention deficit did not show significant differences from patients with neglect (without this deficit) on the FAB test. This may be due to the FAB evaluating a wide range of executive functions, whereas the attention distribution deficit more specifically reflects disturbances in the fronto-parietal network. Impairments in this network may not always be accompanied by significant frontal dysfunction, as measured by standard neuropsychological tests. Additionally, reduced attention to peripheral stimuli and errors in digit identification may be more closely linked to the severity of neglect rather than a separate cognitive deficit, which may also explain the absence of significant differences between the groups on the FAB.

These findings align with results from other studies, supporting the idea that damage to the right temporo-parietal region can disrupt the frontoparietal attention network, impairing the ability to maintain multiple tasks in working memory [58]. Moreover, Bayesian analysis of EEG data has revealed reduced effective connectivity between the right parietal and frontal cortex in patients with neglect, particularly when stimuli appear in the ignored field [87]. Structural neuroimaging studies further emphasize the role of white matter tracts linking the frontal and parietal cortices in spatial attention. Damage to these connections has been associated with more severe forms of neglect and reduced functional connectivity within attention networks [88, 89, 90]. In particular, lesions in the superior longitudinal fasciculus and the supramarginal gyrus have been strongly linked to chronic neglect [91]. These results suggest that non-lateralized attention deficit may be due to impaired fronto-parietal connections rather than primary frontal dysfunction. We suggest that when attention allocation deficits are detected by Keen Eye findings, it is advisable to diagnose impairment of the patient’s executive functions [92]. The impairment of control functions complicates neuropsychological rehabilitation of patients with VN, as it leads to decreased criticality to their condition and difficulties in learning and using voluntary visual search strategies.

Similar effects of impaired attention distribution are also observed in conditions such as acute or chronic mental fatigue, depression, and cognitive aging. For example, studies have shown that moderate mental fatigue reduces the efficiency of attention switching between tasks, which is presumed to be related to the depletion of non-specific cognitive resources [93]. Fatigue has been shown to modulate activity in fronto-parietal and occipital regions, negatively affecting cognitive flexibility [94]. Research indicates that under high cognitive demands, both internal and external attention processes draw from a shared pool of non-specific resources [95]. Similar effects have been described in depression, where excessive rumination and divided attention overload executive control, leading to increased switch costs and impaired divided attention [96]. Likewise, aging is associated with deficits in global task switching, which correlate with reduced working memory capacity, although local switching remains relatively preserved [97]. These examples suggest that the difficulties in attention distribution observed in patients with neglect may result from a broader deficit in cognitive control mechanisms, linked to cognitive resource depletion and extending beyond spatial processing. Understanding these commonalities may provide further insight into the cognitive underpinnings of neglect and aid in developing rehabilitation strategies.

Despite its high diagnostic value, the Keen Eye methodology has some limitations. First, diagnostic accuracy may depend on equipment characteristics, such as screen size and resolution. Furthermore, task performance requires high attention concentration, which may make it difficult to use the methodology with elderly patients. Regarding clinical application, data analysis is conducted using specialized software, requiring staff training. There are also specific visual requirements for patients, as, in clinical practice, it has been difficult for patients to complete the program without glasses. Moreover, the program is unsuitable for patients with severe neurodynamic impairments, as they are unable to complete all 75 trials due to fatigue.

Implementing this methodology into clinical practice could significantly improve neglect diagnosis. Automated data processing can reduce diagnostic time and increase result objectivity. Keen Eye could also be integrated into neurorehabilitation systems to assess patient recovery dynamics after stroke. Furthermore, the methodology could be used for the early detection of cognitive impairments and the monitoring of rehabilitation program efficacy.

For future development of the methodology, several steps are required:

1. Expanding the sample size to include testing with healthy subjects and patients with left hemisphere lesions. This would allow for refinement of diagnostic criteria and increase the specificity of the method.
2. Adapting the methodology for diagnosing neglect in other sensory modalities, such as auditory neglect.
3. Developing mobile versions of the test for convenient use in clinical settings.

The current discussion does not address how specific the methodology is for neglect and how it differentiates neglect from other perception and attention disorders. Future research should include control groups with various visual perception and attention disorders due to brain damage.

## Conclusion

The computerized Keen Eye method has demonstrated high effectiveness in diagnosing neglect, surpassing traditional tests in sensitivity. The study confirmed that the method can detect subtle manifestations of attention impairments and diagnose non-lateralized attention deficits associated with neglect. The dual-task paradigm allows for a comprehensive assessment of attention distribution. By analyzing different indicators, the method can determine the severity of neglect in both horizontal and vertical dimensions. Despite some methodological limitations, Keen Eye has significant potential for clinical application and research, offering an objective, efficient, and adaptable approach for assessing VN and related cognitive disorders.

## Data Availability

All data produced in the present work are contained in the manuscript

## Supporting Information

**S1 File. Database**. This table contains the raw data collected from all study participants using the Keen Eye method and related diagnostic assessments. It includes individual performance metrics such as stimulus omissions, identification errors, and asymmetry coefficients across different conditions and tasks. These data form the basis for the statistical analyses and decision tree models described in the manuscript. Each row corresponds to a single participant, with variables representing key diagnostic indicators.

